# Using qPCR to compare the detection of *Plasmodium vivax* oocysts and sporozoites in *Anopheles farauti* mosquitoes between two DNA extraction methods

**DOI:** 10.1101/2022.11.15.22282365

**Authors:** Lincoln Timinao, Esther W. Jamea, Michelle Katusele, Louis Schofield, Thomas R. Burkot, Stephan Karl

## Abstract

**Background:** Currently, the gold standard to assess parasite developmental stages in mosquitoes is light microscopy. Microscopy can miss low-density infections, is time-consuming and not species-specific. This can place limitations on studies, especially when the infection status of larger mosquito populations is important and studies are done in co-endemic settings with multiple circulating parasite species. Enzyme-linked immunosorbent assay (ELISA) has been the alternative technique to evaluate the infectivity of mosquitoes especially in field studies however it is semi-quantitative. Molecular techniques that have been used to detect the mosquito stages of malaria parasites including *P. vivax*. Here, we present a quantitative real-time assay (qPCR) assay that can be used to detect low-density *P. vivax* oocyst and sporozoite infections. Parasite detection via qPCR after performing the conventional DNA extraction versus direct qPCR following heating of the infected mosquito samples was compared.

**Methods:** Colony reared *Anopheles farauti* mosquitoes were exposed to blood samples collected from infected individuals using a direct membrane feeding assay. The fully fed mosquitoes were kept for 7 and 14 days post-feed before dissection to confirm presence of oocysts and sporozoites. Infected mosquito guts and the salivary glands (with the head and thorax) were stored and DNA was extracted either by heating or by performing conventional column-based DNA extraction. Following DNA extraction the infected samples were subjected to qPCR to detect *P. vivax* parasites.

**Results:** DNA extraction of 1 or more oocysts by heating resulted in an overall sensitivity of 78% (57/73) and single oocysts infections were detected with a sensitivity of 82% (15/17) in the heating arm as well. We observed a 60% (18/30) sensitivity with sporozoites where DNA was extracted using the conventional DNA extraction method prior to qPCR diagnosis. We show that the heating method significantly improved the detection of oocysts over conventional DNA extraction. There was no significant difference in the DNA copy numbers when comparing the detection of oocysts from the conventional DNA extraction versus heating. There was also no significant difference in the detection rate of sporozoite samples when comparing the two DNA extraction protocols. However, we observed that the DNA copy numbers of the sporozoites detected in the heating arm was significantly higher than in the conventional DNA extraction arm.

**Conclusion:** We have adapted a qPCR assay which, when coupled with heating to release DNA reduces sample processing time and cost. Direct qPCR after heating will be a useful tool when investigating transmission blocking vaccines or antimalarials or when evaluating field caught mosquitoes for the presence of malaria parasites.

## Background

Malaria is a significant health problem in 85 countries and nearly half of the world’s population is living in areas with risk of malaria transmission [1]. Despite the efforts to curb malaria globally, it has proven difficult to achieve a steady decrease in malaria cases over the years, highlighting the need for additional interventions. Transmission blocking interventions such as vaccines and antimalarials can be effective tools used to prevent the spread of malaria parasites [2].

Human-to-Mosquito transmission, and the activity of potential transmission-blocking compounds, can be investigated using artificial systems such as membrane feeding set ups. Membrane feeding assays (MFAs) were initially developed by Rutledge and others in the 1960s [3]. In MFAs malaria parasites (whether cultured *in vitro* in the laboratory or from infected patients) are fed to the mosquitoes [4, 5]. Transmission success can be evaluated by the observation of various parasite developmental stages in the mosquito in particular, the oocysts in the midgut and sporozoites in the salivary glands using light microscopy. Traditionally, light microscopy (LM) was used for assessing the presence or absence of the oocysts or sporozoites in the mosquito however, there are inherent limitations with LM detection of parasite mosquito stages. These include labor intensiveness, the requirement for trained personnel and the resulting low throughput. In addition, low-level infections can easily be missed or misdiagnosed, and the differentiation between parasite species in co-endemic settings is not possible.

MFAs can be operationally challenging particularly in resource-limited settings. Since there is no continuous *P. vivax* culture, access to infected individuals is currently the only option. [6] This comes with inherent issues, including in some instances the lack of correlation between the gametocyte densities in natural infections and either the oocyst density or the frequency of mosquito infection. [7, 8] In order to study transmission of malaria parasites derived from infected individuals, a high-throughput method to detect oocysts and sporozoites with high sensitivity is beneficial.

To overcome the limitations of microscopy a number of assays have been developed to enable high throughput detection of parasites in the mosquito gut and salivary glands. These assays include ELISA to detect the circumsporozoite protein (CSP) in mosquito lysates (CSP-ELISA) [9-11], bioluminescence assays to detect transgenic parasites with the green fluorescence protein (GFP) [12-14], near-infrared spectroscopy (NIRS) to detect parasites within mosquitoes [15, 16], enhanced chemiluminescent slot blot (ECL-SB) for detecting *Pf*CSP in mosquito samples [17, 18] and molecular detection of *Plasmodium* DNA [19-21]. Although the CSP-ELISA is relatively robust and cost effective it is only semi quantitative [9-11]. An assay that is quantitative will enable us to know density of the malaria parasites in the mosquito infection. Bioluminescence GFP assays allow for high-through-put but it cannot be used with wild parasites. [12-14] NIRS has been successfully used to detect *P. falciparum* parasites in lab reared mosquitoes with relatively high accuracy but it is still semi quantitative. [15, 16] ECL-SB assays can potentially be used to screen large numbers of mosquitoes for oocysts with high sensitivity and specificity. [17, 18] However, this assay is not quantitative. Various qPCR-based methods have been successfully developed and used to detect blood stage and mosquito infection. However, some qPCR are still semi quantitative mainly due to the design of the qPCR where nonspecific SYBR-green or EVA-green fluorescent dyes were used. [19-22] Taqman assays are an alternative to SYBR-based real time assays. Taqman assays utilise hydrolysis probes that bind to the target sequence and provides a means to quantify the parasite DNA. The Taqman hydrolysis probes have been used to detect blood stage parasites by targeting the 18S ribosomal RNA gene. [23-25] Taqman assays are able to detect parasites at levels 4-5 fold lower than expert thick film microscopy. [26, 27] Taqman assays detect *P. falciparum* [19, 23, 28, 29] and *P. vivax* parasites in mosquitos using minor grove binding (MGB) probes. [28-31] Minor groove binding probes increase the specificity of the probe binding to the target DNA sequence as compared to unmodified probes and limits cross-hybridization of primers and probes in duplexes. [32]

Bass and colleagues established a qPCR assay where they evaluated field caught mosquitoes for the presence of *P. vivax* sporozoites in the head and thorax of individual mosquitoes. They did not investigate the qPCR detection of oocysts or the intensity of sporozoite infections. [28] Rao and colleagues established a multiplex qPCR to detect *Wuchereria bancrofti, P. falciparum*, and *P. vivax* in pools up to 23 field caught mosquitos but did not distinguish between potential sporozoite or oocyst infections. [30] Bickersmith and colleagues also established a qPCR assay on individual field caught mosquitoes but did not distinguish between the oocyst and sporozoite stages as it was not part of the study design. [31] Graumans and colleagues also established a qPCR assay where they successfully detected *P. vivax* oocysts stages in mosquitoes but did not investigate the detection of a single *P. vivax* oocyst as it was not part of the study design. [29] Also they did not investigate the qPCR detection of *P. vivax* sporozoites.

Sample processing time is an important aspect to consider when setting up an MFA or when processing field collected samples. This includes extracting DNA through to qPCR detection of the parasites in the mosquito. DNA extraction using commercially available kits can usually takes several hours depending on the number of samples that are being processed. In a study by Bass and colleagues they heated the mosquito samples for 10 minutes at 95°C and directly performed qPCR after thus reducing the sample processing time. [28] However, they did not evaluate the heating technique against the conventional DNA extraction method. This study addresses the key knowledge gap that exists in setting up a sensitive Taqman qPCR assay for both oocysts and sporozoites with known infection densities and compare the mosquito preparation methods of conventional DNA extraction versus heating.

## Methods

### Mosquito rearing

*Anopheles farauti* mosquitoes were reared at 28 ± 8 ºC and 68 ± 25 % relative humidity (RH) on an 11 h dark and 12 h light including a 30 min dusk and 30 min dawn period. The larvae were fed ground fish food (Marine Master, Tropical Fish Flake) while the adults were provided with 10 % sucrose (Ramu Sugar) solution available as soaked cotton wool balls placed on top of the mosquito cages as previously described. [33] Individuals who provided informed consent performed direct skin feeding to maintain our colony mosquitoes.

### Sample Collection

This study was conducted at the Papua New Guinea Institute of Medical Research (PNGIMR). Ethical approval was received from the PNG Medical Research Advisory Committee (MRAC #16.01). Patients at Yagaum Clinic in Madang Province of PNG, who consented to participate in the study were recruited. Patients were tested with malaria rapid diagnostic tests (RDTs). In the current study the CareStart Malaria Pf/PAN (HRP2/pLDH) Ag Combo RDT kits (Access Bio, Cat No. RMRM-02571CB) were used. Thick and thin blood films were prepared according to WHO methods for evaluation by a certified microscopist. The blood slides were then stained for 30 minutes using 4% Giemsa (Sigma-Aldrich, Australia) stain. [34] Slides were read by the microscopist to identify the presence of the parasites, the species and stages of the parasite in the blood. Parasite density was calculated using the assumption that one microliter of blood contains 8000 white blood cells (WBC) [34] Venous blood samples (5-6mL) were collected from microscopy positive patients in BD Vacutainer ® sampling tubes coated with lithium heparin (BD, Australia). Hemoglobin was measured using a HemoCue® hemoglobin analyzer (HemoCue, Australia). Axillary temperature was taken using a digital thermometer and weight was measured with a bathroom scale (precision ±0.1g). After collection of the blood sample, the BD Vacutainer ® was then immediately stored in a beverage cooler flask (Coleman Company Inc, USA) filled with water adjusted to a temperature of 38°C. A digital thermometer was used to monitor the temperature of the cooler flask. The blood sample was then transported to the insectary for membrane feeding. Transportation time between health facility and laboratory was around 10 minutes.

### Direct membrane feeding assay

At the insectary 3-5 days-old *Anopheles farauti* colony mosquitoes were prepared the previous day and dry starved (i.e., without any sugar or water) overnight. A total of 2 paper cups of 50 mosquitoes per cup were prepared for each feed. Baudruche membrane (Wilco Biotech, USA) was used to feed the mosquitoes through a water-jacketed glass feeder as described previously. [33] Once a blood sample arrived at the insectary it was immediately fed to the mosquitoes for 20 minutes. Unfed mosquitoes were removed and only the fully fed mosquitoes were kept until day 7 post feed when one cup was dissected for oocysts as previously described. [35] The mosquito guts with oocysts were then stored in PBS in Eppendorf tubes at -20°C and then the samples were selected for the thermal treatment and DNA extraction arms. The second cup was held until day 14 post feed for detection of sporozoites. The dissections of salivary glands were done by trained microscopists. The salivary glands that were infected with sporozoites were then stored in 100 - 200µL of PBS buffer together with the head and thorax. The total number of mosquitoes with single oocyst infections together with those with more than one oocyst per mosquito were down-selected for DNA extraction and heating (Appendix 2 Table 1). The sporozoites were classed as low infection (1-20 sporozoites) moderate (21-100) and high (>100) and were also split between the DNA extraction and heating method (Tables S1-S2, Supplementary 1).

**Table 1.** Demographic and clinical data for the study population.

| Demography | Median (range) or<br>n/N (%) |
| --- | --- |
| Age in years (n=68) | 14 (4 - 56) |
| Female (n=68) | 24/68 (35.3) |
| Weight, kg, (n=68) | 40.5 (15 - 77) |
| Haemoglobin, g/dl, (n=65) * | 9.9 (5.6 - 15.5) |
| Temperature, °C , (n=68) | 36.5 (35.3 -39.9) |
| Fever, >37.5 °C, (n=68) | 9/68 (13.2) |

\* Data were not collected for all 68 patients

### Heating and DNA extraction

Parasite DNA was extracted using two methods, the conventional DNA extraction with a commercial kit and heating. In this study we used the FavorPrep® DNA extraction kits (Favorgen Biotech Corp, Ping Tung, Taiwan) and performed DNA extraction according to the protocol for extraction of genomic DNA from tissues and for red blood cells and the DNA was eluted in a final volume of 50 µL of elution buffer. In the heating method, the down-selected samples with oocyst/s and sporozoites were then vortexed for 30 s and then centrifuged for 10 s at 500 g and then heated at 95°C for 10 min.

### Quantitative Real Time PCR (qPCR)

Following heating and DNA extraction of the samples, an established Taqman qPCR assay that utilizes MGB probes was performed to quantify the infection and determine the parasite species. [25] This Taqman qPCR assay was used to detect blood stage parasites. Briefly, this qPCR assay targets the conserved region of the 18SrRNA gene for both *P. falciparum* and *P. vivax*. The quantification of parasite copy numbers is derived from synthetic plasmid DNA of known concentrations that are included in each run. The qPCR was performed on a CFX96 Touch Real-Time Detection System (Bio-Rad, Australia). The primer and probe sequences together with the reaction mix and the thermo profile are shown in Table S1 - S4 in the Supplementary 2 document.

### Statistical Analysis

Statistical analysis was performed using GraphPad Prism (ver. 8.0) and Stata 13 (StataCorp, College Station, TX, USA). The Mann-Whitney test was used to compare the DNA copy numbers between the heating and DNA extraction of mosquito guts with known oocysts counts. The Mann Whitney test was also used to compare the DNA copy numbers between the DNA extraction and the heating method for the sporozoites. The two sample test of proportions was used to compare the proportions of microscopy positive samples that were confirmed by qPCR in the heating and DNA extraction arms.

## Results

A total of 68 patients were recruited (Table 1).

Table 2 shows the results from the three diagnostic methods used.

**Table 2.**
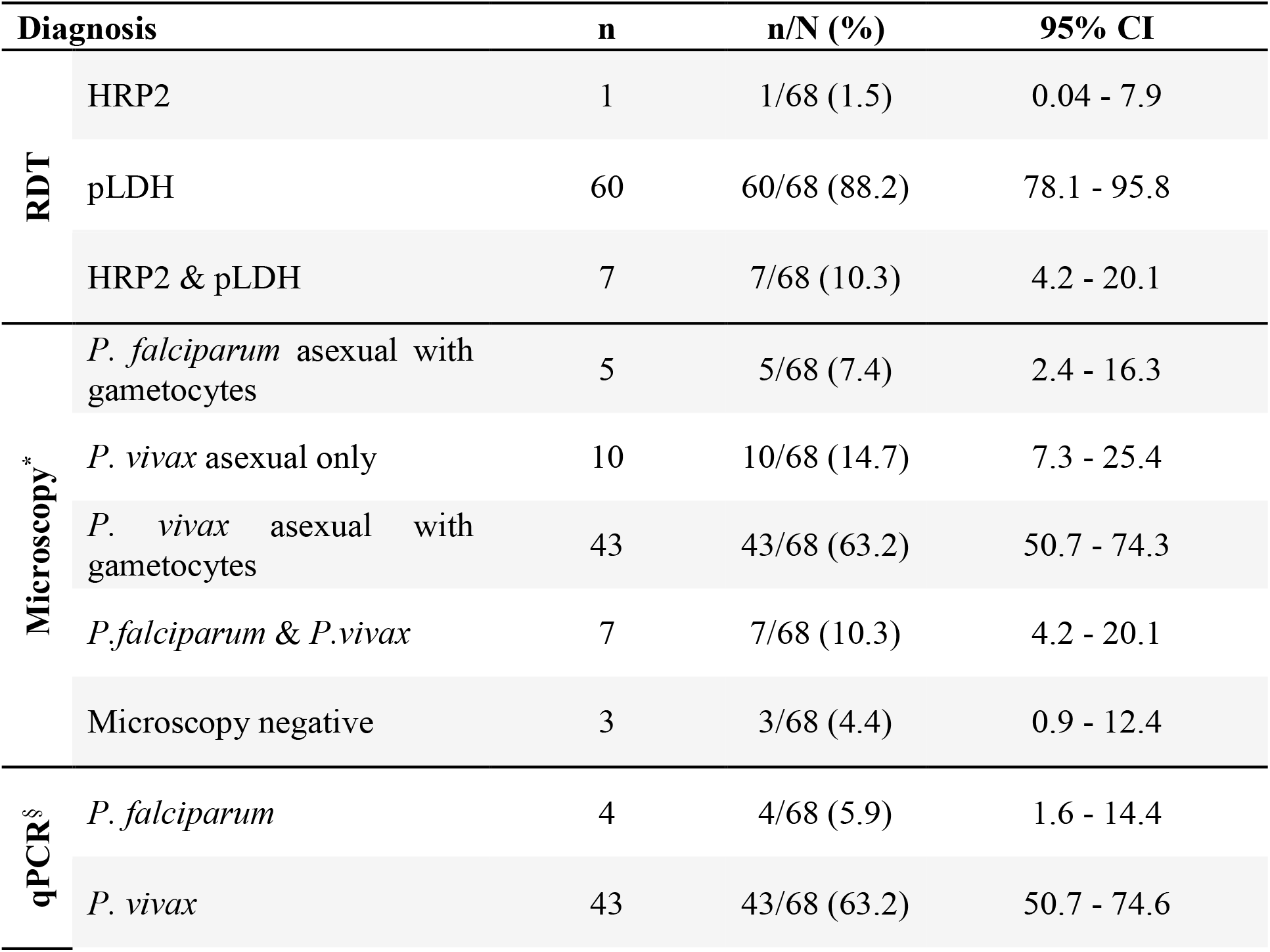
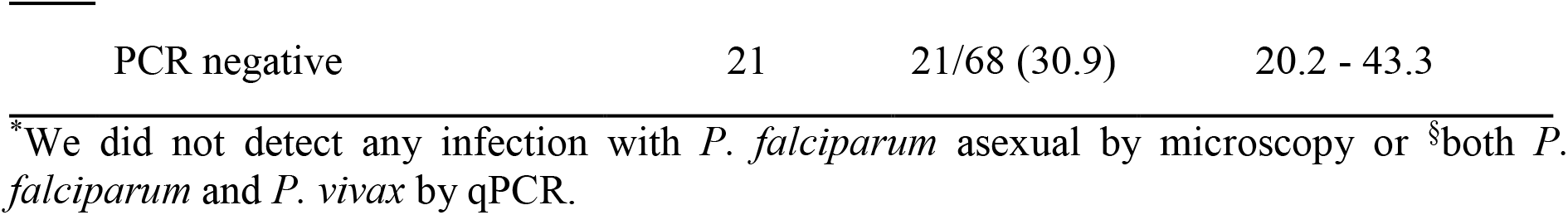
Diagnostic results by RDT, microscopy and qPCR. The number of positive samples per test is n. The total number of samples is N=68. Population averages (n/N(%)) and 95% confidence intervals of proportions (95% CI) are also provided.

We detected *P. vivax* oocyst and sporozoite stages of the malaria parasites in the mosquitoes using our established protocol. Figure 1 shows exemplary amplification curves from a qPCR run.

**Figure 1.**
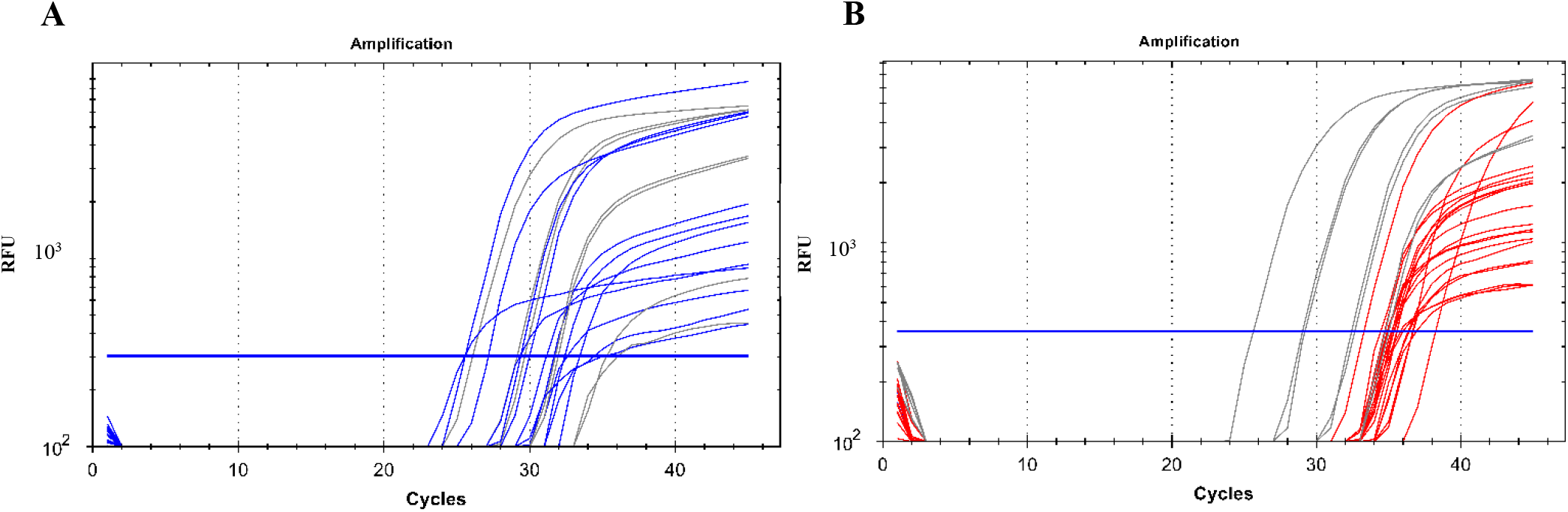
A qPCR amplification plot showing successful amplification of malaria parasite DNA from oocysts and sporozoites. Panel A represents the amplification of parasite DNA from oocysts with the blue curves being the parasite DNA and the grey being the plasmid controls of known concentrations that were used as positive controls. Panel B represents the amplification of parasite DNA from sporozoites with the red curves being the parasite DNA while the grey lines represent the plasmid controls of known concentrations that were used as positive controls. The blue horizontal line represents the threshold value; any curve above this is considered an infection. *RFU - relative fluorescence unit*

A total of 73 and 72 mosquito samples had at least one oocyst in the mosquito gut which was detected by microscopy for the heating and DNA extraction arms respectively. We observed a significantly higher proportion of mosquito samples that were confirmed by qPCR in the heating arm 78% (57/73) as compared to the DNA extraction arm, 39% (28/72) (p<0.0001).

A total of 17 mosquitoes with single oocysts according to microscopy were processed in both the heating and the DNA extraction arm (Table 3). We observed a statistically significant difference with the detection of oocysts by qPCR between the heating arm with a sensitivity of 82% (15/17) and the DNA extraction arm with a sensitivity of 29% (5/17) (p=0.0019).

**Table 3.** Comparison of microscopy positive and qPCR positive oocysts.

|  | Microscopy |  | qPCR |  |
| --- | --- | --- | --- | --- |
|  | No. of mosquitoes | No. of oocysts/mosquito | No. of mosquitoes | Sensitivity %,(n/N) |
| **Heating** | 17 | 1 | 15 | 88 (15/17) |
|  | 5 | 2 | 5 | 100 (5/5) |
|  | 6 | 3 | 4 | 67 (4/6) |
|  | 4 | 4 | 3 | 75 (3/4) |
|  | 2 | 5 | 2 | 100 (2/2) |
|  | 1 | 6 | 0 | 0 |
|  | 1 | 7 | 0 | 0 |
|  | 1 | 10 | 0 | 0 |
|  | 36* | >10 | 33 | 92 (33/36) |
| **DNA Extraction** | 17 | 1 | 5 | 29 (5/17) |
|  | 4 | 2 | 1 | 25 (1/4) |
|  | 4 | 3 | 3 | 75 (3/4) |
|  | 5 | 4 | 2 | 40 (2/5) |
|  | 1 | 5 | 0 | 0 |
|  | 1 | 6 | 0 | 0 |
|  | 1 | 8 | 0 | 0 |
|  | 1 | 13 | 1 | 100 (1/1) |
|  | 1 | 14 | 1 | 100 (1/1) |
|  | 1 | 46 | 1 | 100 (1/1) |
|  | 36* | >10 | 15 | 42 (15/36) |

\*This represents 36 pools of midguts with oocysts and not 36 mosquitoes

When comparing only the oocysts that were successfully detected by qPCR we observed no significant difference between the copy numbers when comparing the detection of parasites from both arms for single oocysts (Figure 2 Panel A). Also there was no significant difference in the DNA copy numbers between the two arms with all mosquitoes with oocysts (Figure 2 Panel B). We also did not observe any correlation with the DNA copy numbers and the oocyst numbers.

**Figure 2.**
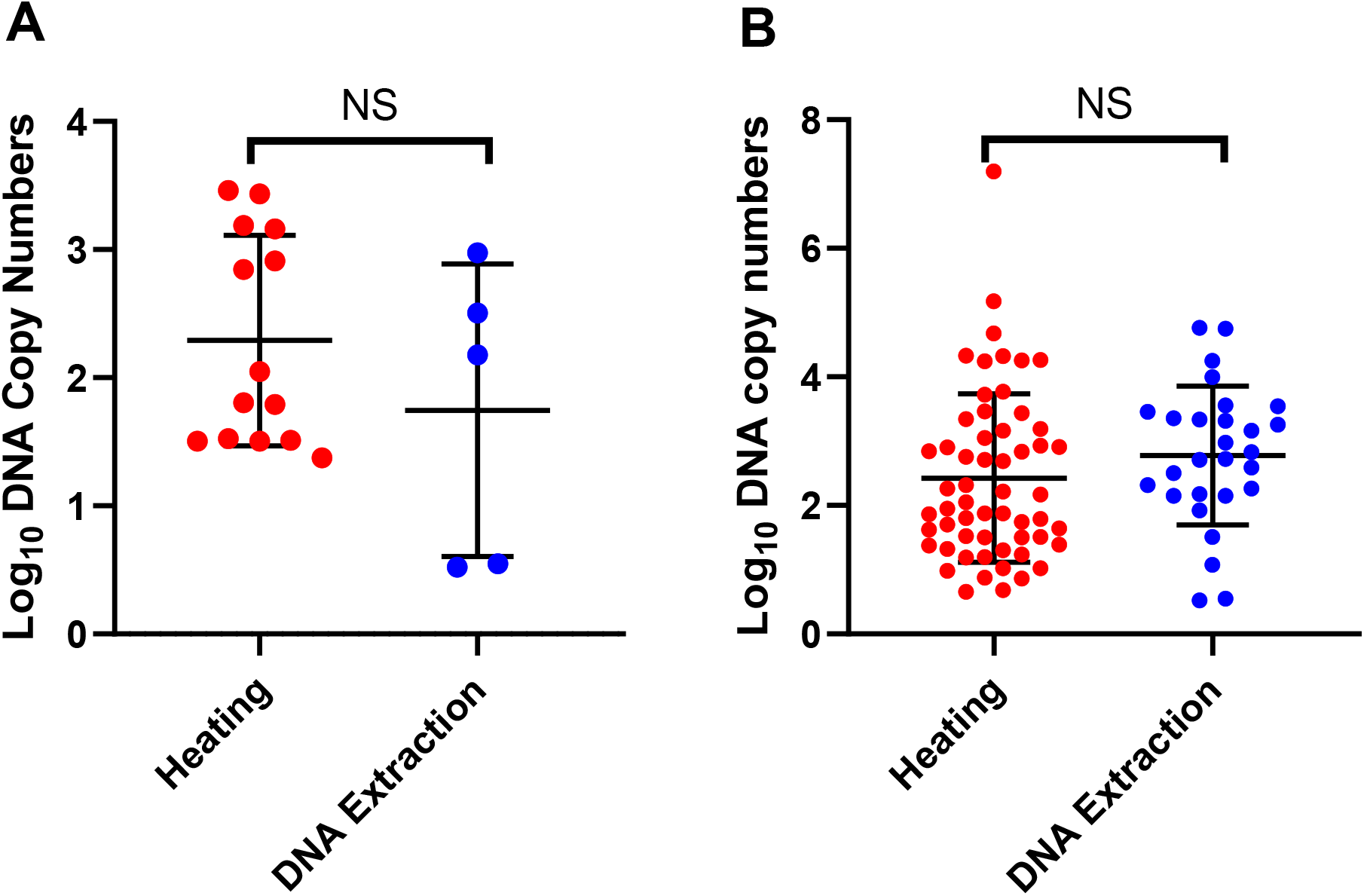
Detection of oocysts using qPCR. Panel A shows the DNA copy numbers from the single oocysts that were detected by qPCR in the two arms. Panel B shows DNA copy numbers of all the mosquito samples with one or more oocysts that were detected by qPCR in the two arms. The error bars show the mean and the standard deviation. The dots are mosquitoes. *NS – Not significant*

A total of 60 mosquito samples positive for sporozoites by microscopy underwent heating (n=30) and DNA extraction (n=30) (Table 4). We observed no significant difference with the detection of sporozoites by qPCR between the heating arm with a sensitivity of 37% (12/30) and the DNA extraction arm with a sensitivity of 60% (18/30) (p=0.121)

**Table 4.**
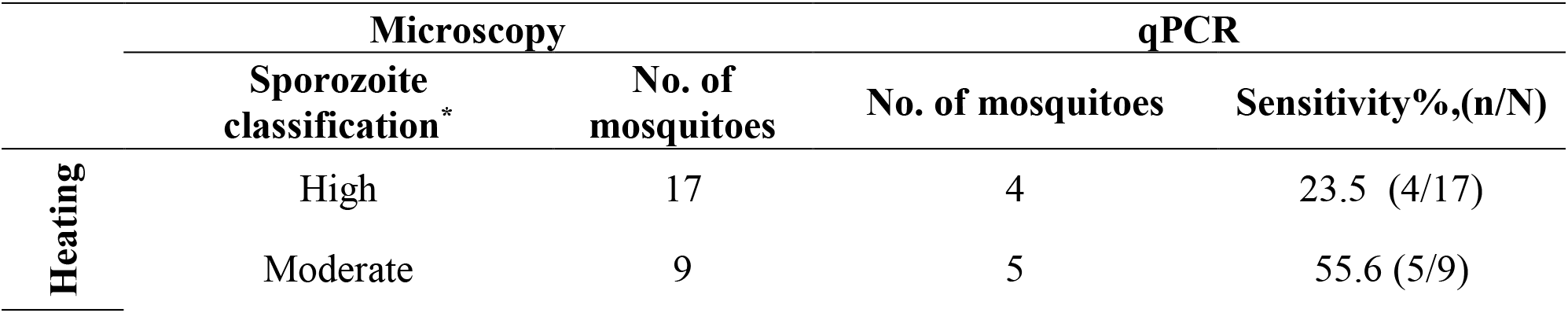
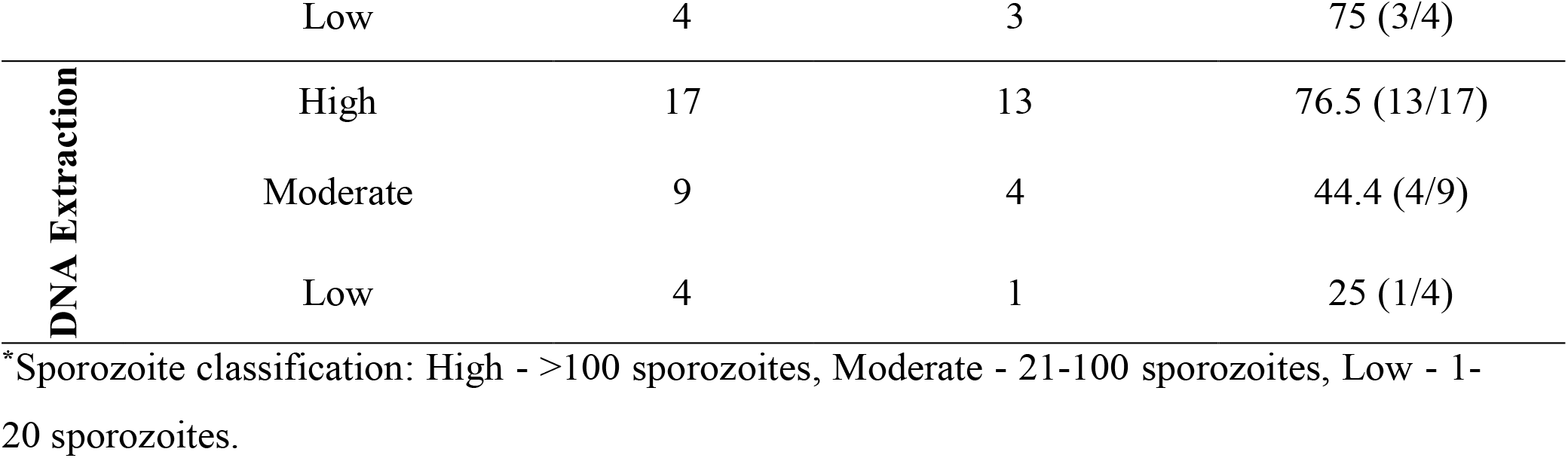
Comparison of microscopy positive and qPCR positive sporozoites.

We observed significantly higher DNA copy numbers (p=0.0126) in the qPCR detection of sporozoites in the heating arm as compared to the DNA extraction arm (Figure 3). We noted that there was a gradual increase in the mean DNA copy number from Low to High sporozoite count (Low: 12.78 (SD, ±19.38), Moderate: 29.85 (SD, ±28.08) and high: 187.29 (SD, ±772.95)).

**Figure 3.**
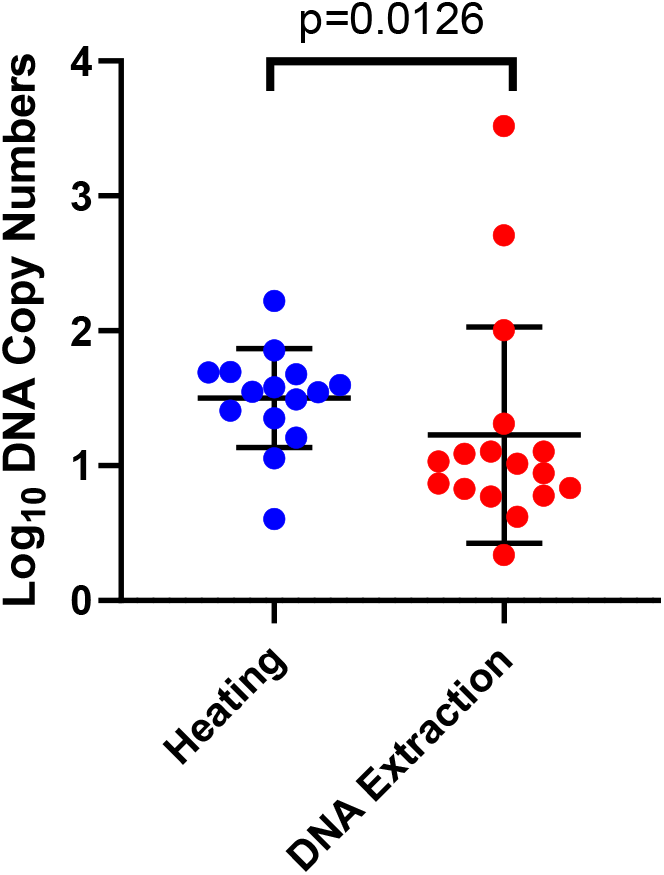
Detection of sporozoites by qPCR in the heating and DNA extraction arms. The error bars show the mean and the standard deviation. Each dot represents a mosquito.

## Discussion

This study describes the adaptation of a high-through-put qPCR based technique for detecting low levels of oocysts and sporozoites and the evaluation of the conventional DNA extraction method versus heating. The qPCR assay is sensitive enough to detect midgut infections with single oocysts. Furthermore, this assay was able to detect low sporozoite infections by microscopy.

Here we have established a qPCR assay that utilizes the Taqman hydrolysis MGB-probe with increased sensitivity in detecting the *P. vivax* parasite target gene and can potentially enable increased through-put for large scale transmission studies. A number of studies have validated TaqMan qPCR assays for detecting *P. vivax* oocysts and/or sporozoites [28-31] but have not investigated the limit of detection. We have shown that this qPCR assay is sensitive in detecting low *P. vivax* oocyst and sporozoites infections in mosquitoes.

We show that there is a higher chance of detecting single oocyst infections when heating the dissected midgut compared to the common method of performing DNA extraction. We also show that there is no significant difference between the detection of the parasite’s DNA copy numbers between heating and DNA extraction especially with low infections indicating that heating has a similar DNA output as the common DNA extraction method. We further observe that there is no significant difference between the DNA copy numbers between the two arms with one or more oocysts.

The current study revealed no significant difference in the qPCR detection of sporozoites between the two techniques used to extract DNA from the microscopy positive salivary glands together with the head and thorax. However, heating yielded significantly higher quantities of DNA copies demonstrating the superior performance of heating over the DNA extraction method.

To our knowledge, this is the first research evaluating heating of mosquito guts and salivary gland (with head and thorax). We show that heating is the better option for releasing oocyst and sporozoite DNA and significantly reduces sample processing time and ensures that samples are processed with high efficiency. It also reduces the cost of processing a sample by skipping DNA extraction step using a conventional DNA extraction kit. Bass and colleagues did use heat to free their *P. falciparum* sporozoite DNA prior to performing qPCR but did not evaluate the sensitivity of the technique. [28] Although similar studies have not been done on mosquitoes, we found that similar comparisons were made with bacteria where they evaluated heating the samples versus using commercially available DNA extraction kits. They found no significant difference between the PCR output from both techniques and suggested that heating was efficient, simple, cheap and suitable for high-through-put.[36, 37] Similar to what was seen in the case of bacteria, heating the mosquito midguts and salivary glands yielded similar qPCR detection rates for sporozoites while higher detection rates with oocysts as compared to DNA extraction.

## Conclusion

In summary, we show that a qPCR assay can be used to detect very low numbers of mosquito stage *P. vivax* parasites. Furthermore, we show that by heating the mosquito guts and the head and thorax we save on costs and reduce the time taken to process the samples. We believe that this high-through-put setup will be a valuable tool in evaluating potential transmission blocking vaccines or antimalarials or for evaluating the infection status of field caught mosquitoes.

## Data Availability

All data produced in the present study are available upon reasonable request to the authors

