## Supplementary Tables for "Using qPCR to compare the detection of *Plasmodium vivax* oocysts and sporozoites in *Anopheles farauti* mosquitoes between two DNA extraction methods"

Supplementary 1

**Table 1.** Oocyst counts and whether they were processed by heating or DNA extraction.

| Number of oocyst/mosquito | Number of Mosquitoes | Heating / DNA extraction |
|---|---|---|
| 1 | 17 | Heating |
| 2 | 5 | Heating |
| 3 | 6 | Heating |
| 4 | 4 | Heating |
| 5 | 2 | Heating |
| 6 | 1 | Heating |
| 7 | 1 | Heating |
| 10 | 1 | Heating |
| Pools | 36 | Heating |
| 1 | 17 | DNA extraction |
| 2 | 4 | DNA extraction |
| 3 | 4 | DNA extraction |
| 4 | 5 | DNA extraction |
| 6 | 1 | DNA extraction |
| 8 | 1 | DNA extraction |
| 13 | 1 | DNA extraction |
| 14 | 1 | DNA extraction |
| 46 | 1 | DNA extraction |
| Pools | 36 | DNA extraction |

**Table 2.** Sporozoites classification and whether they were processed by heating or DNA extraction.

| Sporozoite classification* | Number of Mosquitoes | Heating / DNA extraction |
|---|---|---|
| High | 17 | Heating |
| Moderate | 9 | Heating |
| Low | 4 | Heating |
| Negative | 8 | Heating |
| High | 17 | DNA extraction |
| Moderate | 9 | DNA extraction |

| | | |
|---|---|---|
| Low | 4 | DNA extraction |
| Negative | 8 | DNA extraction |

* High >100 sporozoites, Moderate 20 -100, Low 1-20 sporozoites

**Supplementary 2**

**Table 1.** Primer sequences of for the qPCR assay to detect *P. falciparum* and *P. vivax* parasites

| Species | Primer[1] | Sequence (5' - 3') |
|---|---|---|
| ***P. falcipaurm*** | **Pf_fwd** | TATTGCTTTTGAGAGGTTTTGTTACTTTG |
| | **Pf_rev** | ACCTCTGACATCTGAATACGAATGC |
| ***P. vivax*** | **Pv_fwd** | GCTTTGTAATTGGAATGATGGGAAT |
| | **Pv_rev** | ATGCGCACAAAGTCGATACGAAG |

**Table 2.** Probe sequences for the qPCR assay to detect *P. falciparum* and *P. vivax* parasites.

| Species | Probe[2] | Sequence (5' - 3') |
|---|---|---|
| ***P. falciparum*** | **Pf probe** | **6FAM**-ACGGGTAGTCATGATTGAGTT-**MGBNFQ** |
| ***P. vivax*** | **Pv probe** | **VIC**-AGCAACGCTTCTAGCTTA -**MGBNFQ** |

**Table 3.** The reaction mix for the qPCR

| qPCR Reaction mix |
|---|
| **Total volume** 14µL |
| 2X Roche Master mix[3] |
| 350nM per primer ( forward and reverse) |
| 350nM per probe ( forward and reverse) |
| 4µL of DNA |

**Table 4.** The cycling conditions for the qPCR.

| Thermo profile | | | |
|---|---|---|---|
| **Hold** | 50°C | 2min | |
| **Hold** | 95°C | 15min | |
| **Denaturation** | 95°C | 15sec | X 45 |
| **Annealing** | 60°C | 1min | |

[1.] Integrated DNA Technologies (IDT), New Zealand
[2.] LifeScience Roche, NSW, Australia
[3.] TaqMan MGB Probes ThermoFisher Scientific, Auckland, New Zealand
